# A Machine Learning Ensemble Based on Radiomics to Predict BI-RADS Category and Reduce the Biopsy Rate of Ultrasound-Detected Suspicious Breast Masses

**DOI:** 10.1101/2021.12.16.21267907

**Authors:** Matteo Interlenghi, Christian Salvatore, Veronica Magni, Gabriele Caldara, Elia Schiavon, Andrea Cozzi, Simone Schiaffino, Luca Alessandro Carbonaro, Isabella Castiglioni, Francesco Sardanelli

## Abstract

We developed a machine learning model based on radiomics to predict the BI-RADS category of ultrasound-detected suspicious breast lesions and support medical decision making towards short-interval follow-up versus tissue sampling. From a retrospective 2015–2019 series of ultrasound-guided core needle biopsies performed by four board-certified breast radiologists using six ultrasound systems from three vendors, we collected 821 images of 834 suspicious breast masses from 819 patients, 404 malignant and 430 benign according to histopathology. A balanced image set of biopsy-proven benign (n = 299) and malignant (n = 299) lesions were used for training and cross-validation of ensembles of machine learning algorithms supervised during learning by histopathological diagnosis as a reference standard. Based on a majority vote (over 80% of the votes to have a valid prediction of benign lesion), an ensemble of support vector machines showed an ability to reduce the biopsy rate of benign lesions by 15% to 18%, always keeping a sensitivity over 94%, when externally tested on 236 images from two image sets: 1) 123 lesions (51 malignant and 72 benign) obtained from the same four ultrasound systems used for training, resulting into a positive predictive value (PPV) of 45.9% (95% confidence interval 36.3-55.7%) versus a radiologists’ PPV of 41.5% (*p* < 0.005), combined with a 98.0% sensitivity (89.6–99.9%); 2) 113 lesions (54 malignant and 59 benign) obtained from two ultrasound systems from vendors different from those used for training, resulting into a 50.5% PPV (40.4–60.6%) versus a radiologists’ PPV of 47.8% (*p* < 0.005), combined with a 94.4% sensitivity (84.6–98.8%). Errors in BI-RADS 3 category (i.e., assigned by the model as BI-RADS 4) were 0.8% and 2.7% in the *Testing set I* and *II*, respectively. The board-certified breast radiologist accepted the BI-RADS classes assigned by the model in 114 masses (92.7%) and modified the BI-RADS classes of 9 breast masses (7.3%). In 6 of 9 cases the model performed better than the radiologist, since it assigned a BI-RADS 3 classification to histopathology-confirmed benign masses that were classified as BI-RADS 4 by the radiologist.

## 1. Introduction

Ultrasound imaging is a key tool in breast care. Indications to breast ultrasound, recently summarized by the European Society of Breast Imaging (EUSOBI) [1], include: palpable lump; axillary adenopathy; first approach for clinical abnormalities in women younger than 40 years of age and in pregnant or lactating women; suspicious abnormalities revealed at mammography or contrast-enhanced magnetic resonance imaging (MRI); suspicious nipple discharge; skin retraction; recent nipple inversion; breast inflammation; abnormalities at the site of intervention after breast conserving surgery or mastectomy; abnormalities in the presence of oncoplastic or aesthetic breast implants. Moreover, when MRI is not performed, the following indications to breast ultrasound can be considered: screening high-risk women or women with extremely dense breasts (supplemental to mammography); loco-regional staging of a known breast cancer; monitoring breast cancers receiving neoadjuvant systemic therapy. In addition, ultrasound provides an optimal, cheap, and comfortable guidance for performing needle biopsy for suspicious ultrasound-detected breast lesions, including those initially detected at digital mammography techniques (two-dimensional, tomosynthesis, or contrast-enhanced mammography) or MRI, when a sure correlation between the ultrasound finding and the initially detected finding can be established [2,3].

Indeed, since benign abnormalities (and sometimes also normal breast tissues) are able to mimic malignancies even on advanced breast imaging modalities and techniques, tissue sampling represents the best method for confirmation or exclusion of breast cancer [2,3]. Thus, in the last decades, percutaneous needle biopsy has been established as a crucial approach to prevent unnecessary surgery, reduce associated morbidity as well as economic and psychological costs associated with suspicious findings finally being demonstrated to be benign. The European Society of Breast Cancer Specialists (EUSOMA) includes, among the mandatory quality indicators in breast cancer care [4], the assessment of the “proportion of women with breast cancer (invasive or in situ) who had a preoperative histologically or cytologically confirmed malignant diagnosis (B5 or C5)”. For this indicator, EUSOMA requires a “minimum standard” rate of 85% and a target rate of 90% [4]. The dark side of the moon of the worldwide practice of percutaneous breast needle biopsy, mostly performed under ultrasound guidance, is the variable and frequently high rate of procedures needed to exclude malignancy for finding that finally are revealed to be benign. To avoid missing cancers, breast radiologists are “forced” to biopsy also many abnormalities with probably benign features, unless they think that a given lesion in a given patient, also considering patient-specific risk factors (family and personal history as well as clinical conditions), has an extremely low probability of being malignant and that a six-month delayed diagnosis will not impact on patient’s outcome. Using the Breast Imaging Reporting and Data System (BI-RADS), this means to categorize the lesion as BI-RADS 3, which should imply a residual cancer probability lower than 2%, against a cancer probability higher than 2% but lower than 95% (BI-RADS 4) and a cancer probability higher than 95% (BI-RADS 5) [5]. New approaches aiming at reducing the ultrasound-guided biopsy rate of benign breast lesions must take into account such a challenging clinical context.

Machine learning is a methodological approach of artificial intelligence that concerns building systems that learn based on the data they use. It is widely used in medical imaging to develop image-driven multivariate systems effective in complex tasks, such as supporting physicians in clinical decision making [6]. Radiomics, i.e., the measurement of a high number of quantitative features from images characterizing size, shape, image intensity and texture of identified findings, has been extensively used to train multivariate machine learning algorithms to objectively characterize image findings and to predict diagnosis and prognosis of individual lesions or subjects. In breast cancer care, radiomics has been applied to a variety of medical image modalities for the aforementioned purposes, including mammography, digital breast tomosynthesis, ultrasound, magnetic resonance imaging, and positron-emission tomography combined with computed tomography [7–10], with good performances and with the advantage of high explainability, in particular when the radiomic predictors of the models can be compared and interpreted with reference to semantic predictors previously described in literature. In particular, many features of breast lesions on ultrasound images are known to be associated with higher or lower probability of malignancy of a given lesion, as Stavros et al. [11] pointed out in their seminal paper focused on breast solid masses published more than 25 years ago. These authors described traditional features such as shape, margins, spatial orientation, absolute signal intensity, signal intensity relative to the surrounding tissue (the classic hyper-, iso-, and hypoechoic patterns), and signal heterogeneity, all of them integrated in the BI-RADS lexicon [5]. However, quantification and integration of such a wide spectrum of information is difficult to be attained by a human reader, whereas it is expected to be best achieved through a multivariate model of radiomics and machine learning.

Therefore, the aim of our study was to develop and validate a machine learning model based on radiomics to classify ultrasound-detected suspicious breast masses with the specific two-fold purpose of providing a second-opinion on BI-RADS classification and of reducing the needle biopsy rate. A high sensitivity combined with a sizable reduction in the number of false positive cases was the guiding criteria to develop the machine learning model. The best radiomic predictors were specifically described and interpreted to explain the model and its results.

## 2. Materials and Methods

This study retrospectively analyzed the breast biopsy database of the Radiology Unit at IRCCS Policlinico San Donato (San Donato Milanese, Milan, Italy) and was approved by the institutional ethics committee (Comitato Etico IRCCS Ospedale San Raffaele, protocol code “SenoRetro”, first approved on November 9^th^, 2017, then amended on July 18^th^, 2019, and on May 12^th^, 2021). The acquisition of specific informed consent was waived due the retrospective nature of the study.

### 2.1. Study population and image sets

A consecutive series of 926 patients referred for ultrasound-guided core-needle biopsy from January 13^th^, 2014, to May 28^th^, 2019, was retrieved, for a total of 928 ultrasound images of 941 suspicious breast masses according to the judgment of one of four rotating certified breast radiologists with 4 to 14 years of experience in breast imaging. All ultrasound images were acquired with one of six ultrasound systems (Esaote MyLab 6100, MyLab 6150, MyLab 6440, and MyLab 7340002, Esaote S.p.A, Genova, Italy; Samsung RS80A, Samsung Healthcare, Seoul, South Korea; Acuson Juniper, Siemens Healthineers, Erlangen, Germany). After database search, another certified breast radiologist with 34 years of experience in breast imaging retrospectively reviewed all images to identify the biopsied lesion on the ultrasound images, excluding 96 images from 96 women for which a sure identification of the biopsied mass was not attainable. Ultimately, 821 ultrasound images of 834 suspicious breast masses from 819 patients (mean age 56 ± 16 [standard deviation] years) were considered for radiomic analysis and to develop and test the machine learning model. Histopathology from core needle biopsy or pathology of surgical specimens was used as a reference standard, with 404/834 lesions (48.4%) proven to be malignant and 430/834 lesions (51.6%) proven to be benign, for an overall 1.06:1.00 benign-to-malignant ratio.

A balanced set of randomly sampled ultrasound images from 299 malignant and 299 benign lesions, all from four of the six ultrasound systems (Esaote MyLab 6100, MyLab 6150, MyLab 6440, and MyLab 7340002), were used for the training and internal testing of different ensembles of machine learning classifiers, based on the supervised learning of histopathology as a reference standard (*Training and internal testing set*). Then, the remaining images of 123 other lesions (51 malignant and 72 benign according to histopathology) obtained from the same four ultrasound systems of the *Training and internal testing set*, were used as first external testing for the best machine learning model (*Testing set I*). Finally, the remaining images of the 113 lesions (54 malignant and 59 benign according to histopathology), obtained from the other two of the six considered ultrasound systems (Samsung RS80A and Siemens Healthineers Acuson Juniper), were used as second external testing for the best machine learning model (*Testing set II*).

### 2.2. Radiomic-based machine learning modelling

Radiomic methodology was applied to the 821 included images, according to the International Biomarker Standardization Initiative (IBSI) guidelines [12]. For this purpose, the TRACE4© radiomic platform [13] was used allowing the whole IBSI-compliant radiomic workflow to be obtained in a fully-automated way. The IBSI radiomic workflow included: *i*) segmentation of the suspicious mass to obtain a region of interest (ROI) from each patient image; *ii*) preprocessing of image intensities within the segmented ROI required to measure radiomic features; *iii*) measurement of radiomic features from the segmented ROI; *iv*) the use of such candidate radiomic features to train, validate, and test different models of machine learning classifiers in the binary classification task of interest (malignant *versus* benign discrimination), by the reduction of such extracted features to reliable and nonredundant features.

More specifically, in this study:

1. The segmentation of suspicious masses on all 821 images was performed manually by a board-certified radiologist with 34 years of experience in breast imaging, using the TRACE4 segmentation tool. The same radiologist (at a time distance of 8 weeks) and a second board-certified radiologist with 7 years of experience, independently segmented the masses on a random subsample of 50 images from the training dataset, fully blinded to histopathology and other segmentations.
2. The preprocessing of image intensities within the segmented ROI included resampling to isotropic voxel spacing, using a downsampling scheme by considering an image slice thickness equal to pixel spacing, and intensity discretization using a fixed number of 64 bins.
3. The radiomics features measured from the segmented ROI were 107 quantitative descriptors and belonged to different families: morphology, intensity-based statistics, intensity histogram, gray-level co-occurrence matrix (GLCM), Gray-Level Run Length Matrix (GLRLM), Gray-Level Size Zone Matrix (GLSZM), Neighborhood Gray Tone Difference Matrix (NGTDM), Gray-Level Distance Zone Matrix (GLDZM), Neighboring Gray Level Dependence Matrix (NGLDM). Their definition, computation and nomenclature are compliant with the IBSI guidelines, except for the features of the family morphology, originally designed for 3D images, which were replaced with ten 2D equivalent features (e.g. 3D features volume and surface were replaced with 2D features area and perimeter, respectively). Radiomic features were selected as those showing an intraclass correlation coefficient > 0.75 among the two intra-observer and inter-observer segmentations on the random subsample of images described in 1). Steps from 2) to 3) were performed using the TRACE4 Radiomics tool. Radiomic features were reported by TRACE4 according to IBSI standards.
4. Three different models of machine learning classifiers were trained, validated, and tested, for the binary classification task of interest (malignant *versus* benign discrimination), based on supervised learning, using histopathology as a reference standard. For each model, a nested k-fold cross validation method was used (k = 10, 8 folds for training, 1 fold for tuning, 1 fold for hold-out testing, random sampling). The first model consisted of 3 ensembles of 100 random-forest classifiers combined with Gini index with majority-vote rule; the second model consisted of 3 ensembles of 100 support vector machines combined with principal components analysis and Fisher discriminant ratio with majority-vote rule; the third ensemble consisted of 3 ensembles of 100 k-nearest neighbor classifiers combined with principal components analysis and Fisher discriminant ratio with majority-vote rule. The performances of the 3 models were measured across the different folds (k = 10) in terms of sensitivity, specificity, area under the receiver operating characteristic curve (ROC-AUC), positive predictive value (PPV), negative predictive value (NPV) and corresponding 95% confidence intervals (CI). The model with the best performance according to ROC-AUC was chosen as the best classification model for the binary task of interest (malignant *versus* benign discrimination).

A study of the percentage of the votes of the classifiers in an ensemble to have a valid prediction (concordance on predicted class higher than a qualified majority) of benign and malignant lesions was performed during cross validation in order to maximize sensitivity. Ultimately, this machine learning model was tested on the two external datasets (*Testing set I* and *Testing set II*).

Relevant radiomic predictors were selected as those radiomic features most frequently chosen by the machine learning classifiers as the most relevant ones during the cross validation of the ensembles. For random forest classifiers, the mean importance of each radiomic feature was obtained by each random forest classifier during validation on out-of-bag samples. For support vector machines and k-nearest neighbor classifiers, the mean weight coefficient of each radiomic feature was obtained as explained by each principal component selected by the classifier through a grid search on validation samples.

### 2.3. BI-RADS diagnostic categories classification

When the percentage of the votes of the classifiers in the best ensemble had a valid prediction of benign lesions (concordance on predicted benign class higher than the qualified majority), the ensemble assigned the BI-RADS 3 category. Similarly, when the percentage of the votes of the classifiers in the best ensemble had a valid prediction of malignant lesions, the ensemble assigned the BI-RADS 4 or 5 category according to the level of concordance of the majority of support vectors in the ensemble. For each of the breast masses of the *Testing set I* (123 masses), the certified breast radiologist with 34 years of experience in breast imaging accepted or modified the BI-RADS category assigned by the best ensemble (best model), blinded to histopathology. The class agreement and disagreement were assessed on a case by case basis using histopathology as reference standard. Of course, in this assessment, BI-RADS categories 1 (no abnormalities), 2 (benign lesions), 0 (inconclusive examination), and 6 (known malignancy) were not considered due to the design of the study. The class agreement and disagreement of the random subsample of images, resegmented by the board-certified radiologist with 34 years of experience (intra-observer agreement) and by the board-certified radiologist with 7 years of experience (inter-observer agreement), were assessed on a case by case basis using the first segmentation of the board-certified radiologist with 34 years of experience as reference standard. For each comparison between reference standard segmentation and the two resegmentations, mean DICE indices were obtained. Also for this assessment, BI-RADS categories 1 (no abnormalities), 2 (benign lesions), 0 (inconclusive examination), and 6 (known malignancy) were not considered due to the design of the study.

### 2.4. Statistical analysis

Statistical analysis was conducted with embedded tools of the TRACE4 platform. To describe the distribution of each of the most relevant features in the malignant and benign classes we calculated their medians with 95% CIs and presented violin and box plots.

A non-parametric univariate Wilcoxon rank-sum test (Mann-Whitney *U* test [14]) was performed for each of the relevant radiomic predictors to verify its significance in discriminating malignant from benign lesions. To account for multiple comparisons, the *p* values were adjusted using the Bonferroni-Holm method and the significance levels were set at 0.05 (*) and 0.005 (**) [15].

## 3. Results

### 3.1. Study population and image sets

Table 1 details the histopathological classification of the 834 suspicious breast lesions included in the study, while Table 2 lists technical information about the acquisition of the 821 ultrasound images that depicted these 834 lesions and their distribution into image sets used for all phases of the machine learning model development. A total of six different ultrasound systems were considered, four from the same vendor, the other two from different vendors, with an overall mean image pixel size ranging from 0.062 mm to 0.106 mm. The study population comprised 13 males and 806 females, aged 56.0 ± 16.1 years (mean ± standard deviation).

**Table 1.**
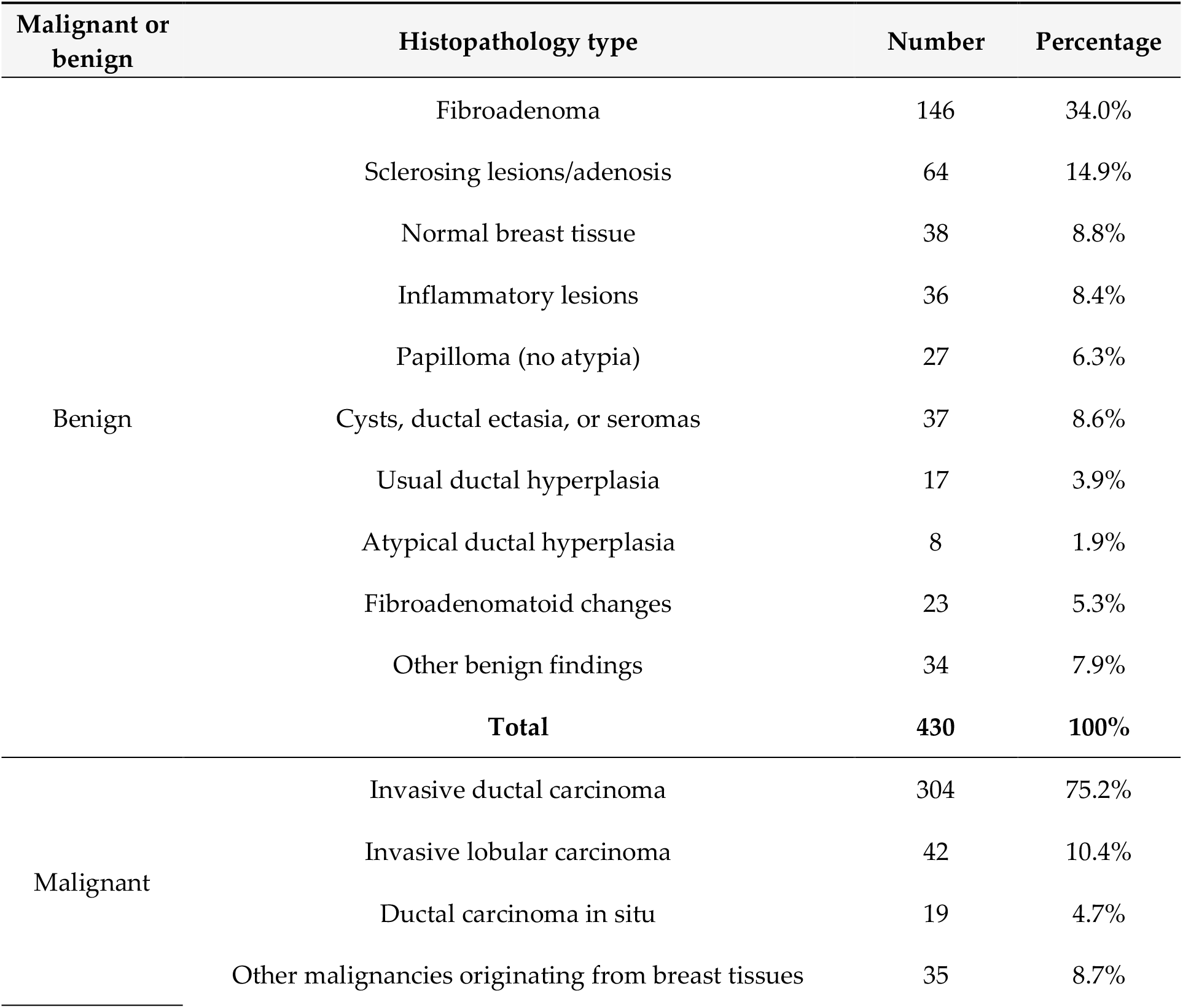
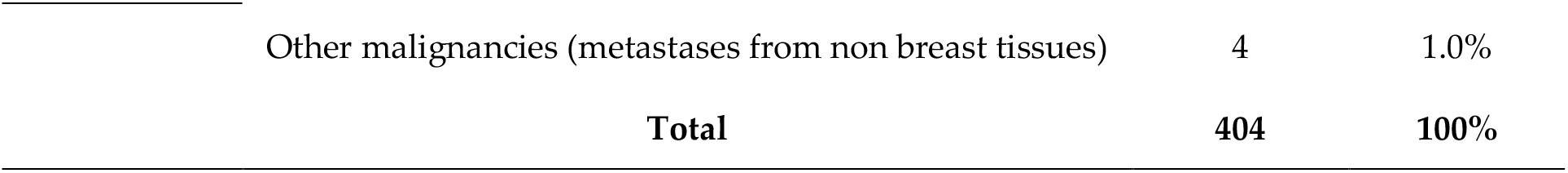
Histopathology of the 834 breast masses included in the study.

| Malignant or benign | Histopathology type | Number | Percentage |
| --- | --- | --- | --- |
| Benign | Fibroadenoma | 146 | 34.0% |
|  | Sclerosing lesions/adenosis | 64 | 14.9% |
|  | Normal breast tissue | 38 | 8.8% |
|  | Inflammatory lesions | 36 | 8.4% |
|  | Papilloma (no atypia) | 27 | 6.3% |
|  | Cysts, ductal ectasia, or seromas | 37 | 8.6% |
|  | Usual ductal hyperplasia | 17 | 3.9% |
|  | Atypical ductal hyperplasia | 8 | 1.9% |
|  | Fibroadenomatoid changes | 23 | 5.3% |
|  | Other benign findings | 34 | 7.9% |
|  | **Total** | **430** | **100%** |
| Malignant | Invasive ductal carcinoma | 304 | 75.2% |
|  | Invasive lobular carcinoma | 42 | 10.4% |
|  | Ductal carcinoma in situ | 19 | 4.7% |
|  | Other malignancies originating from breast tissues | 35 | 8.7% |

|  |  |  |
| --- | --- | --- |
| Other malignancies (metastases from non breast tissues) | 4 | 1.0% |
| **Total** | **404** | **100%** |

**Table 2.**
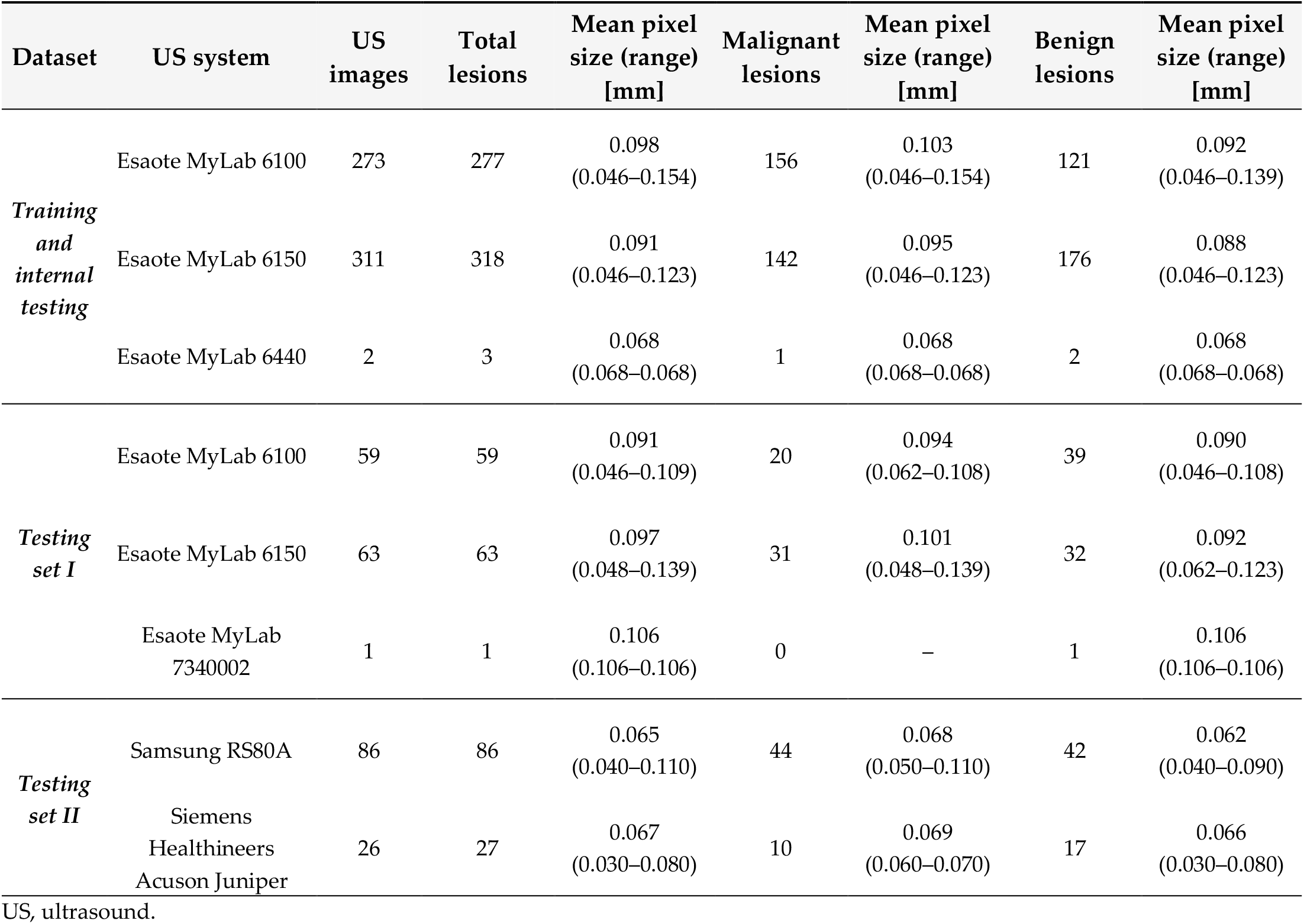
Technical details and composition of the three image sets.

### 3.2. Radiomic-based machine learning modelling

A total of 107 radiomic features were found stable among the two intra-observer and inter-observer segmentations on the random subsample of images and were used to train (nested k-fold cross validation) and externally test the machine learning ensembles.

The ensemble of support vector machines resulted to be the best system for the task of interest, i.e., discrimination between biopsy-proven benign *versus* malignant lesions, performance comparison for all ensembles being shown in Table S1, Table S2, and Table S3. A majority vote > 80% of machines to have a valid prediction of benign lesions and a majority vote > 50% of machines to have a valid prediction of malignant lesions warranted a sensitivity > 94% during both training and external testing, that is the crucial performance to be warranted for ultrasound examination of suspicious breast lesions (Figure 1), allowing however a reduction of 15– 18% in the number of the needle biopsies which resulted in benign histopathology; this consensus was chosen as a qualified majority vote for the task of interest in this specific clinical context. Interestingly, as depicted in Figure 1, the sensitivity was > 96% on images from different ultrasound systems but from the same vendor (*Training and internal testing set* and *Testing set I*).

**Figure 1.**
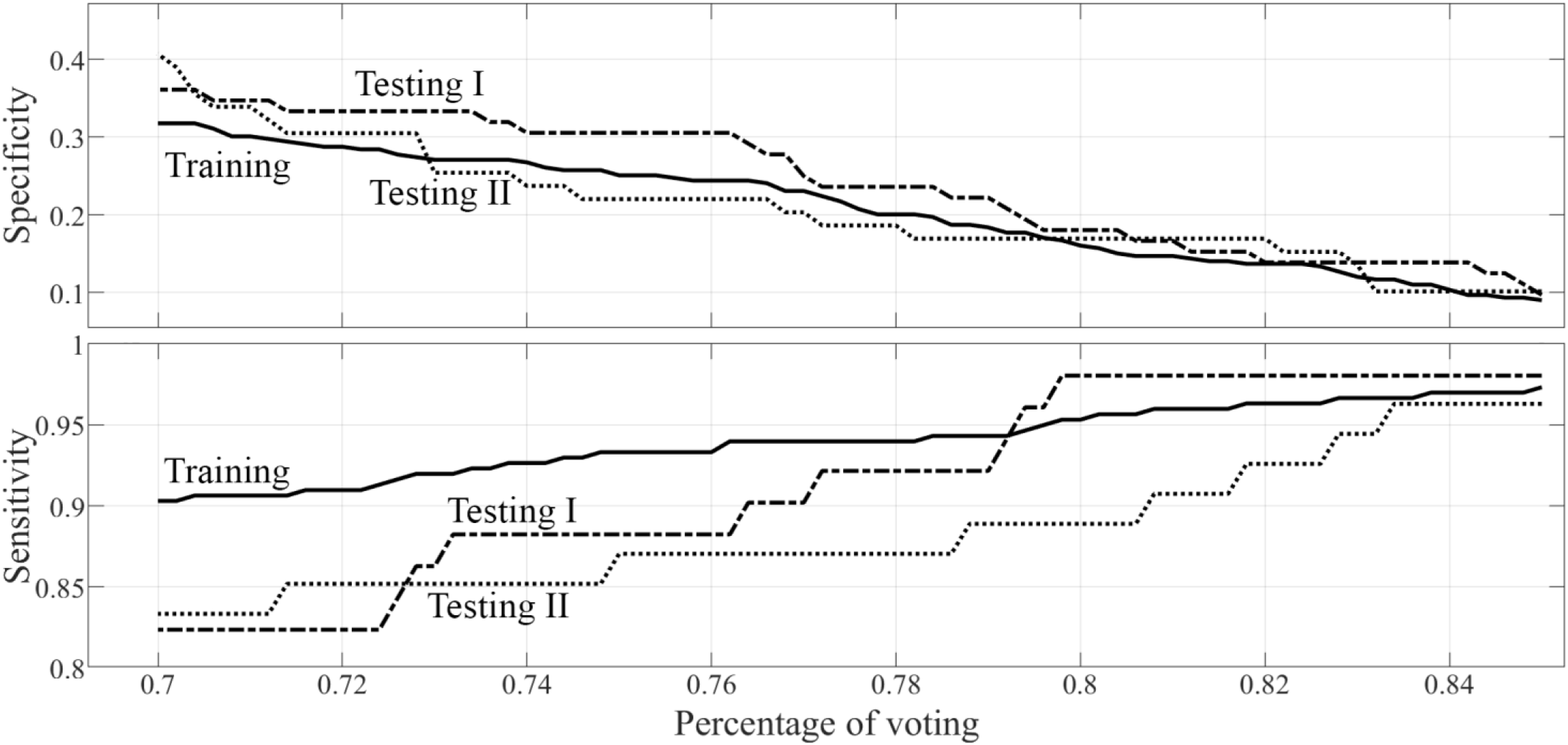
Ensemble of support vector machines: proportion of correctly predicted benign and malignant lesions *versus* percentage of voting from the support vector machines.

Performance metrics of this high-sensitivity machine learning model in the *Training and internal testing set* (10 fold cross-validation) were: sensitivity 95.7%** (95% CI 92.7–97.7%), NPV 78.3%** (65.8–87.9%), PPV 53.2%** (48.8–57.4%), and specificity 15.7%** (11.8–20.3%). As detailed in Table 3, also presenting comparisons of PPV and specificity with those achieved by the radiologists’, performances of the machine learning model in the external *Testing set I* were: sensitivity 98.0% (89.6–99.9%), NPV of 92.9% (66.1–99.8%), PPV 45.9%** (36.3–55.7%), and specificity 18.1** (10.0–28.9%). Performances in the external *Testing set II* were sensitivity 94.4% (84.6– 98.8%), NPV of 75.0% (42.8–94.5%), PPV of 50.5%** (40.4–60.6%), and specificity 15.3%** (7.2–27.0%).

**Table 3.**
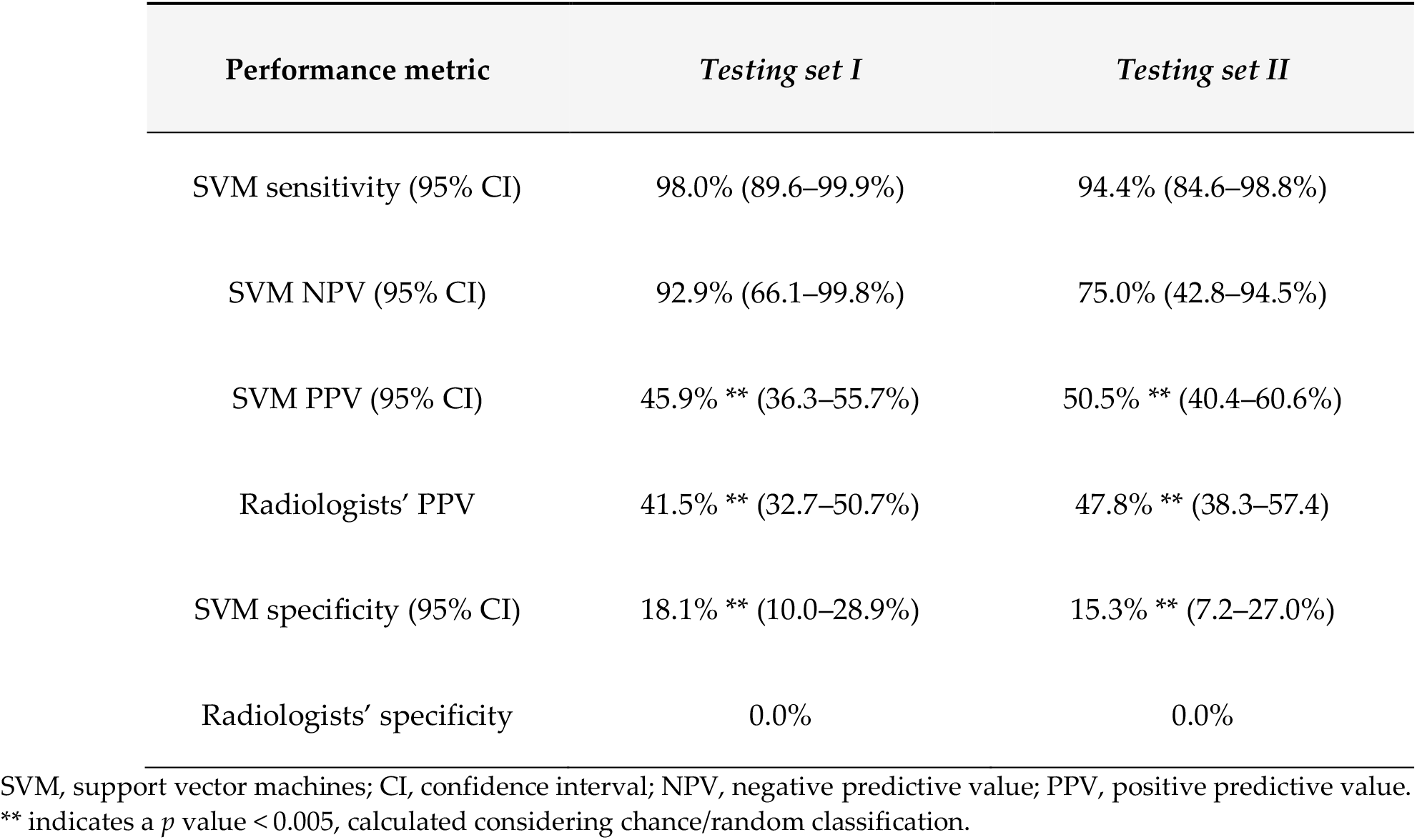
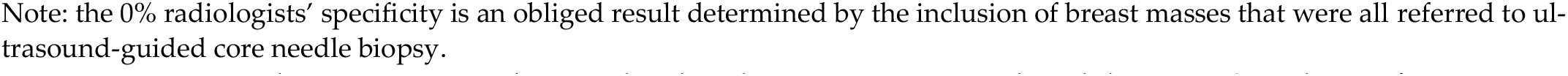
Performances of the ensemble of support vector machines in the external testing datasets.

| Performance metric | Testing set I | Testing set II |
| --- | --- | --- |
| SVM sensitivity (95% CI) | 98.0% (89.6–99.9%) | 94.4% (84.6–98.8%) |
| SVM NPV (95% CI) | 92.9% (66.1–99.8%) | 75.0% (42.8–94.5%) |
| SVM PPV (95% CI) | 45.9% ** (36.3–55.7%) | 50.5% ** (40.4–60.6%) |
| Radiologists' PPV | 41.5% ** (32.7–50.7%) | 47.8% ** (38.3–57.4) |
| SVM specificity (95% CI) | 18.1% ** (10.0–28.9%) | 15.3% ** (7.2–27.0%) |
| Radiologists' specificity | 0.0% | 0.0% |

SVM, support vector machines; CI, confidence interval; NPV, negative predictive value; PPV, positive predictive value.
\*\* indicates a $p$ value $< 0.005$ , calculated considering chance/random classification.
Note: the 0% radiologists' specificity is an obliged result determined by the inclusion of breast masses that were all referred to ultrasound-guided core needle biopsy.

Principal components analysis and Fisher discriminant ratio reduced the 107 IBSI-radiomic features, measured from each breast lesion of the *Training and internal testing set*, to an average of 12 (range 7™17) independent principal components for each support vector machine of the ensemble. The top-25 most relevant radiomic predictors selected by such model from the 107 IBSI-compliant features are shown in Table 4, together with their IBSI feature family and feature nomenclature, and ranked according to their frequencies among the most relevant ones in the support vector machines of the ensemble. Results from univariate statistical sum rank tests are also reported with adjusted *p* values. The violin plots and boxplots of the first 15 radiomic predictors are shown in Figure 2, while the violin plots and boxplots of the other 10 radiomic predictors are shown in Figure S1.

**Table 4.** Ensemble of support vector machines. Top-25 most relevant predictors sorted in descending order of relevance.

| Rank | Feature family | Feature name |
| --- | --- | --- |
| 1 | Morphology | Perimeter to area ratio** |
| 2 | Morphology | Maximum diameter** |
| 3 | Morphology | Compactness** |
| 4 | Morphology | Acircularity** |
| 5 | Morphology | Perimeter** |
| 6 | Morphology | Area** |
| 7 | Morphology | Center of mass shift** |
| 8 | Morphology | Circularity* |
| 9 | Neighborhood grey tone difference matrix | Strength** |
| 10 | Neighborhood grey tone difference matrix | Coarseness** |
| 11 | Neighborhood grey tone difference matrix | Contrast |
| 12 | Neighborhood grey tone difference matrix | Busyness* |
| 13 | Grey-level size zone matrix | Zone size non uniformity** |
| 14 | Grey-level size zone matrix | Grey level non uniformity glszm** |
| 15 | Neighboring grey level dependence matrix | Dependence count non uniformity** |
| 16 | Neighboring grey level dependence matrix | Low dependence low grey level emphasis |
| 17 | Grey-level run length matrix | Grey level non uniformity |
| 18 | Grey-level run length matrix | Run length non uniformity |
| 19 | Intensity-based statistics | Minimum |
| 20 | Intensity-based statistics | Energy |
| 21 | Intensity-based statistics | Variance |
| 22 | Intensity-based statistics | Quartile coefficient |
| 23 | Intensity-based statistics | 10<sup>th</sup> percentile |
| 24 | Intensity histogram | 10<sup>th</sup> percentile |
| 25 | Grey-level co-occurrence matrix | First measure of information correlation |

\* denotes statistical significance at 0.05 (adjusted with Bonferroni-Holm correction). \*\* denotes a statistical significance at 0.005 (adjusted with Bonferroni-Holm correction).

**Figure 2.**
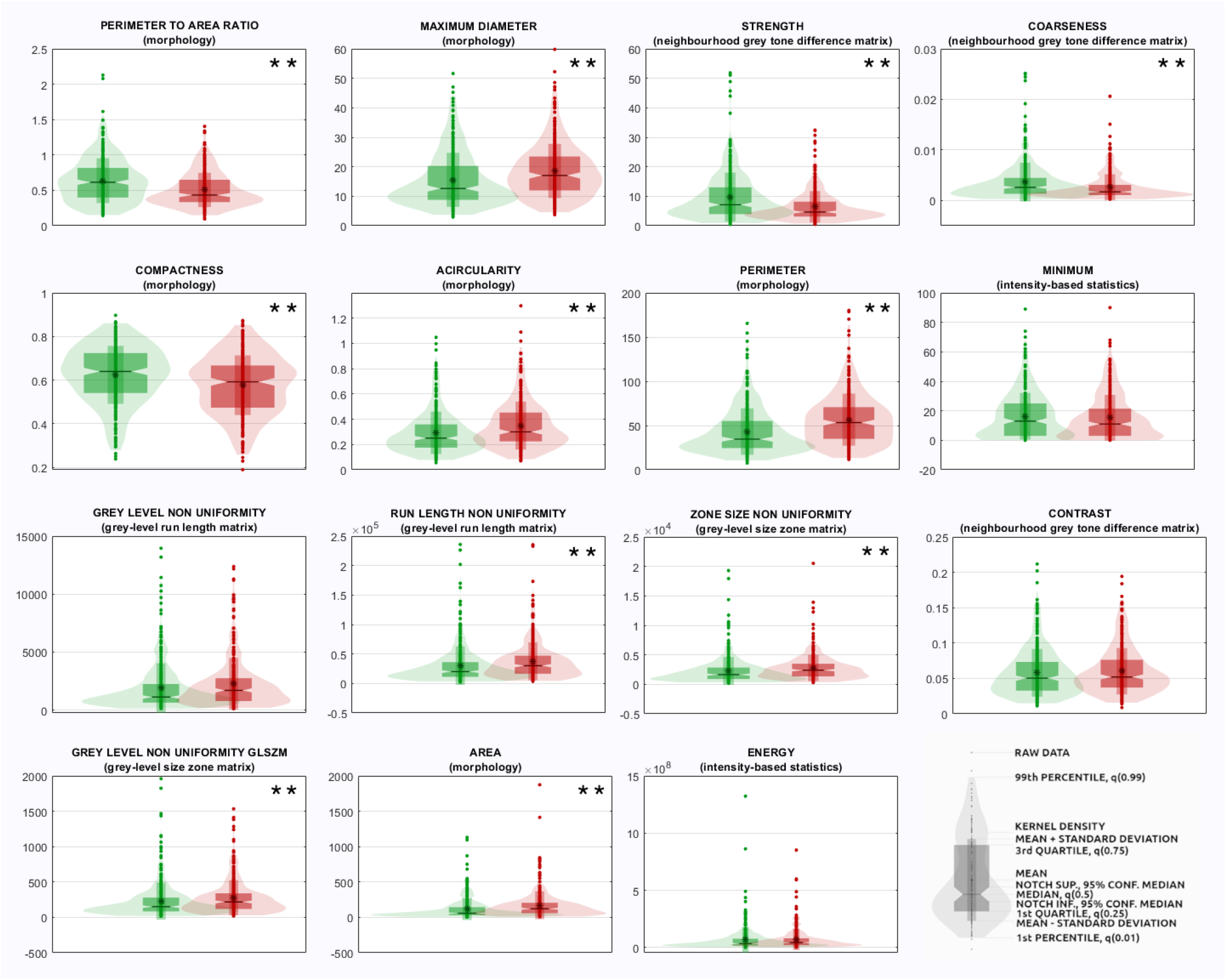
Ensemble of support vector machines: Violin and box plots of the most relevant radiomic predictors ranked from 1 to 15. Green: benign class. Red: malignant class. *denotes statistical significance at 0.05 (adjusted with Bonferroni-Holm correction). ** denotes a statistical significance at 0.005 (adjusted with Bonferroni-Holm correction).

Figure 3 and Figure 4 depict examples of breast masses according to histological diagnosis as classified by the developed radiomic-based machine learning system. ROIs manually defined by the expert breast radiologist to segment the suspicious lesion are overlapped on the corresponding images. The 107 measured IBSI-compliant radiomic features are reported for these lesions in Table S4.

**Figure 3.**
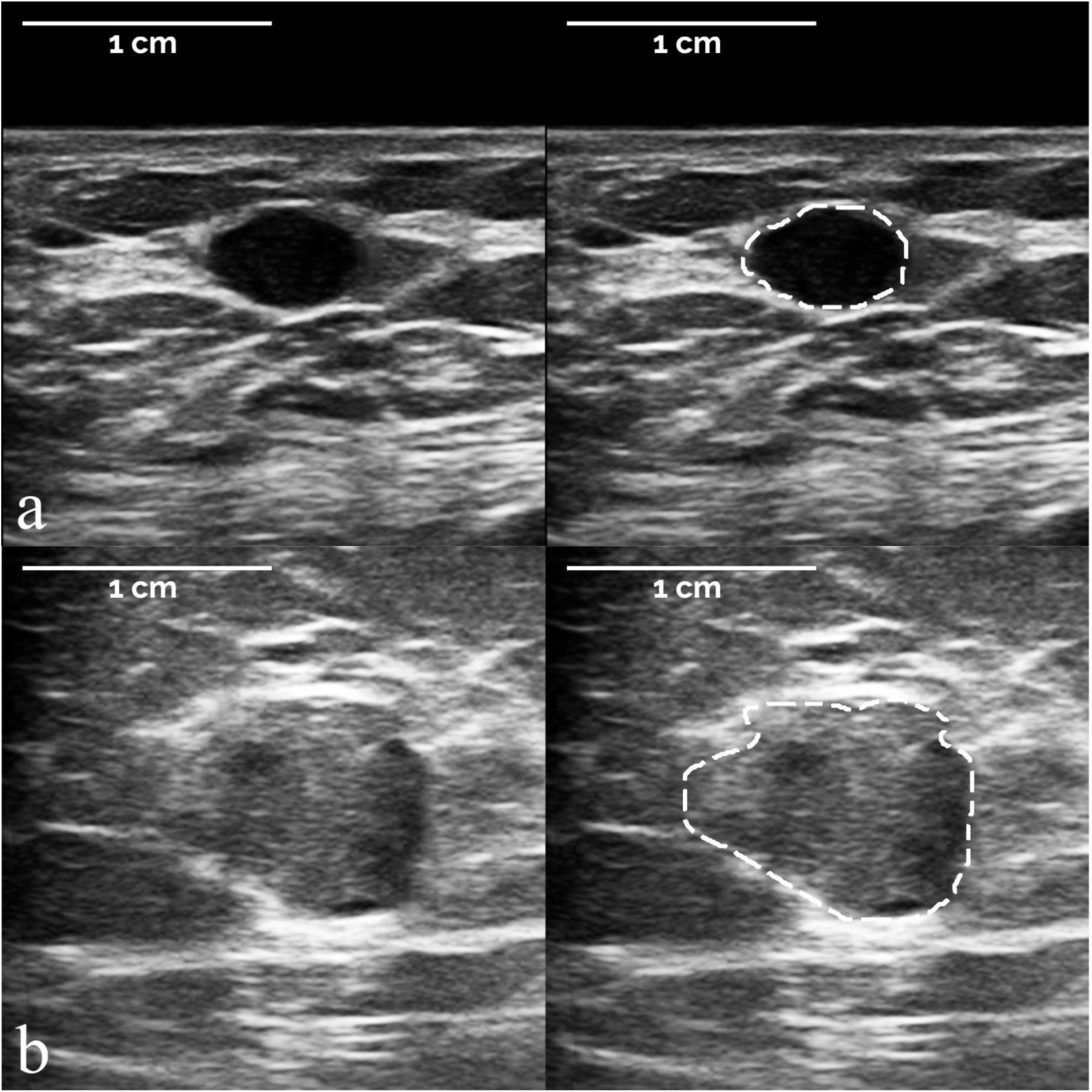
Representative examples of two benign lesions according to histological diagnosis, as classified by the developed radiomic machine learning system. First row (**a**): true negative (benign lesion correctly classified as < 2% likelihood of cancer); second row (**b**): false positive (benign lesion incorrectly classified as > 2% likelihood of cancer). ROIs were manually defined by the expert breast radiologist to segment the suspicious breast lesion. Radiomic features: (**a**) *compactness* 0.807; *acircularity* 0.113; *center of mass shift* 3.495; *zone size non-uniformity* 743.6. (**b**) *compactness* 0.718; *acircularity* 0.181; *center of mass shift* 4.458; *zone size non-uniformity* 2255.5. Histopathology: (**a**) cyst; (**b**) fibroepithelial proliferation.

**Figure 4.**
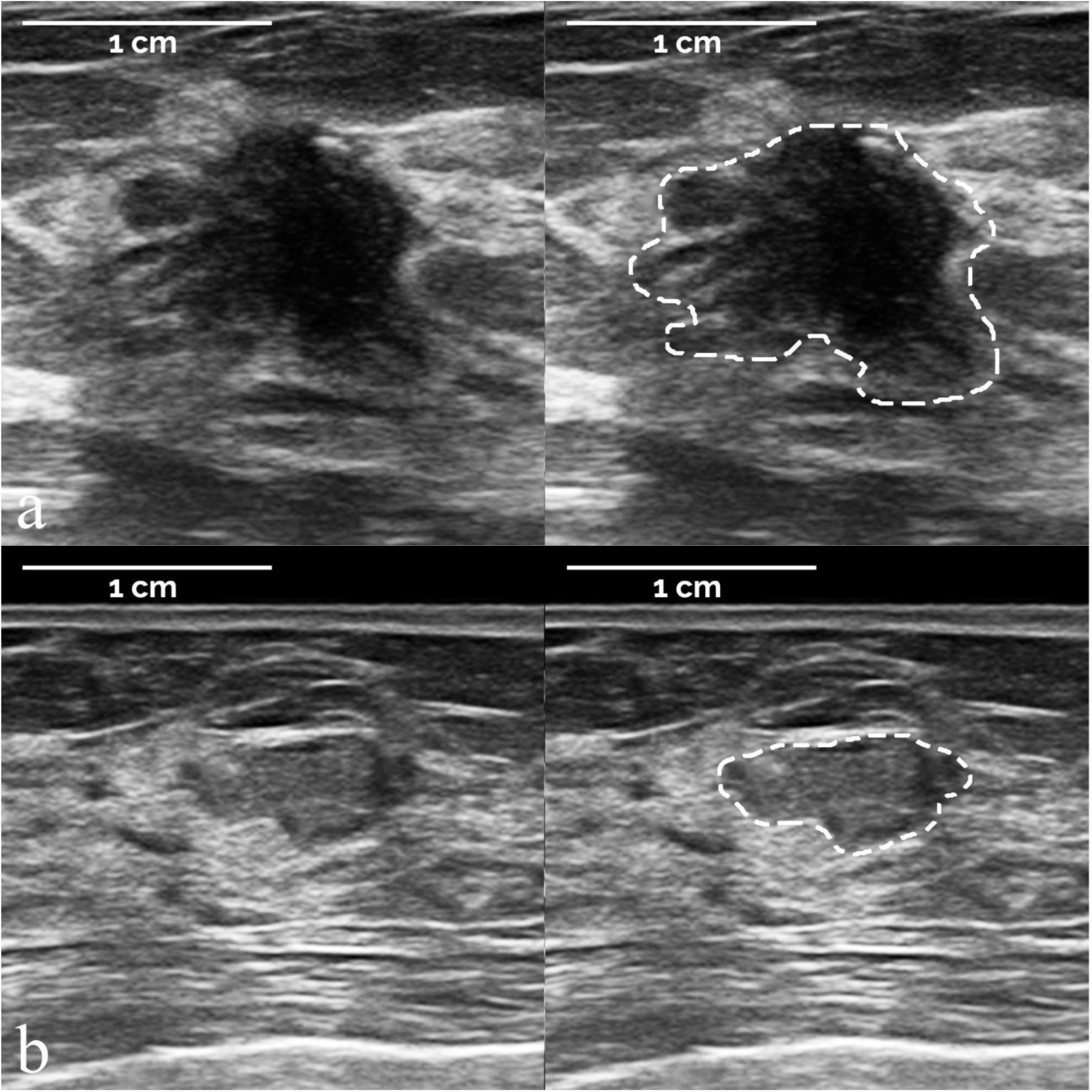
Representative examples of two malignant lesions according to histological diagnosis, as classified by the developed radiomic machine learning system. First row (**a**): true positive (malignant lesion correctly classified as > 2% likelihood of cancer); second row (**b**): false negative (malignant lesion incorrectly classified as < 2% likelihood of cancer).= ROIs were manually defined by the expert breast radiologist to segment the suspicious breast lesion. Radiomic features: (**a**) *compactness* 0.569; *acircularity* 0.326; *center of mass shift* 3.997; *zone size non-uniformity* 2600.2. (**b**) *compactness* 0.614; *acircularity* 0.276; *center of mass shift* 5.861; *zone size non-uniformity* 2209.0. Histopathology: (**a**) invasive ductal carcinoma; (**b**) papillary carcinoma.

### 3.3. BI-RADS diagnostic categories classification

Table 5, Table 6, and Table 7 show the distribution of the BI-RADS categories with respect to histopathology groups as assigned by the ensemble of support vector machines to the breast masses of the *Training and internal testing set* (598 masses), *Testing set I* (123 masses), and *Testing set II* (113 masses). Errors in BI-RADS 3 category assignments by the model were 0.8% and 2.7% in *Testing set I* and *Testing set II*, respectively.

**Table 5.** Ensemble of support vector machines: BI-RADS diagnostic categories predicted for breast masses of the *Training and internal testing set* according to histopathology groups.

| Histopathology type | BI-RADS 3 (%) | BI-RADS 4 (%) | BI-RADS 5 (%) |
| --- | --- | --- | --- |
| Fibroadenoma | 8 (1.3) | 93 (15.6) | 0 (0.0) |
| Sclerosing lesions/adenosis | 10 (1.7) | 37 (6.2) | 2 (0.3) |
| Normal breast tissue | 6 (1.0) | 26 (4.3) | 1 (0.2) |
| Inflammatory lesions | 2 (0.3) | 26 (4.3) | 0 (0.0) |
| Papilloma (no atypia) | 1 (0.2) | 17 (2.8) | 0 (0.0) |
| Cysts, ductal ectasia, or seromas | 10 (1.7) | 13 (2.2) | 0 (0.0) |
| Usual ductal hyperplasia | 2 (0.3) | 9 (1.5) | 0 (0.0) |
| Atypical ductal hyperplasia | 0 (0.0) | 0 (0.0) | 0 (0.0) |
| Fibroadenomatoid changes | 2 (0.3) | 13 (2.2) | 0 (0.0) |
| Other benign findings | 8 (1.3) | 93 (15.6) | 0 (0.0) |
| Invasive ductal carcinoma | 5 (0.8) | 211 (35.3) | 6 (1.0) |
| Invasive lobular carcinoma | 2 (0.3) | 32 (5.4) | 1 (0.2) |
| Ductal carcinoma in situ | 4 (0.7) | 10 (1.7) | 0 (0.0) |
| Other malignancies originating from breast tissues | 2 (0.3) | 24 (4.0) | 0 (0.0) |
| Other malignancies (metastases from non breast tissues) | 0 (0.0) | 1 (0.2) | 1 (0.2) |

**Table 6.**
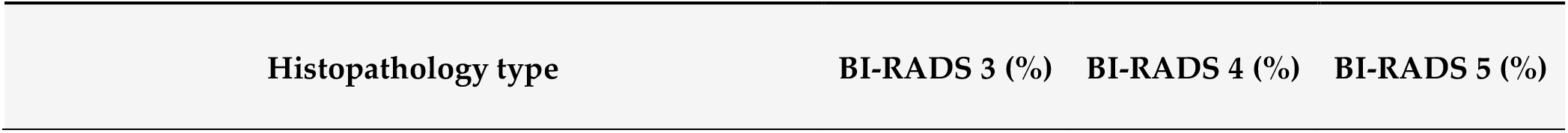
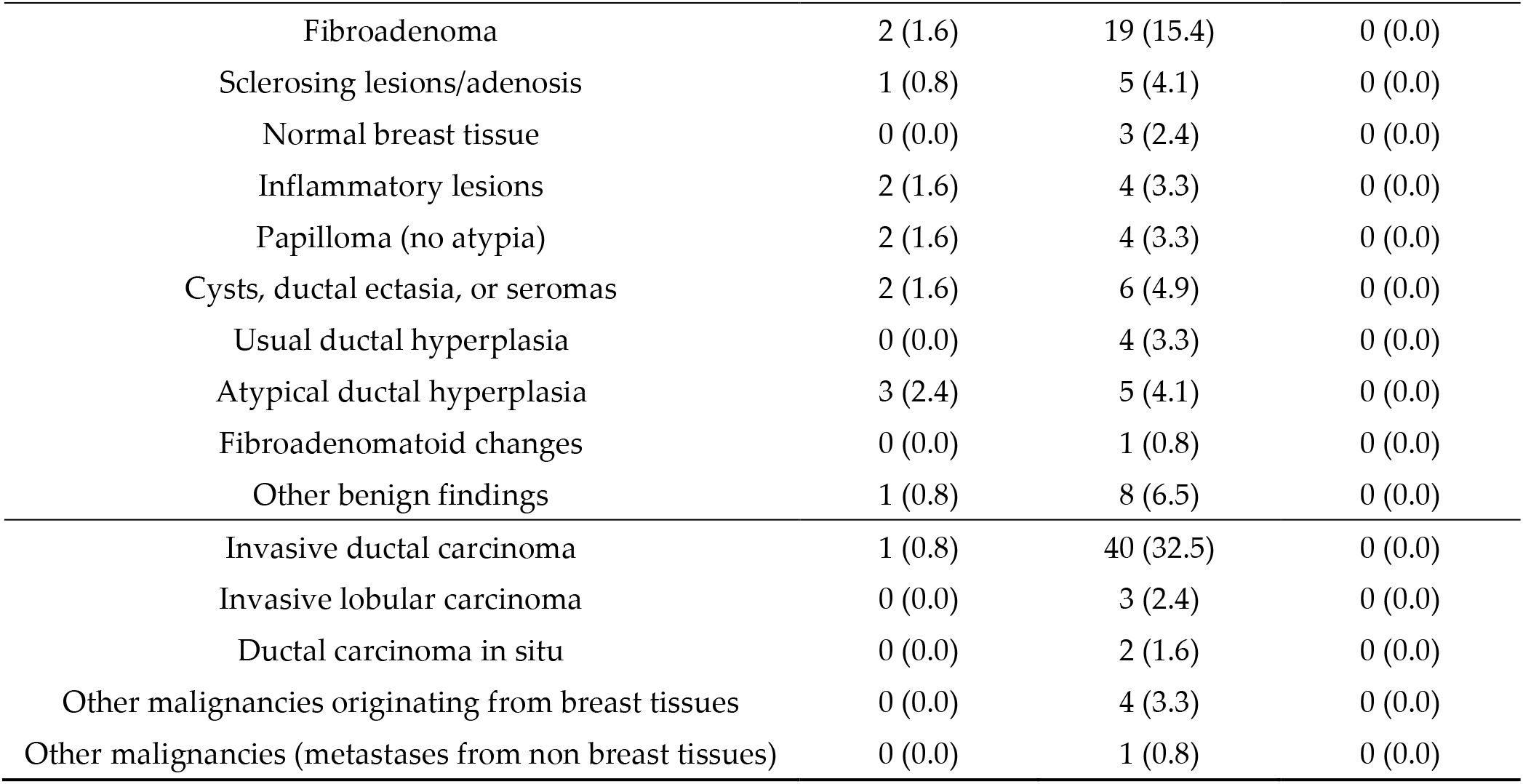
Ensemble of support vector machines: BI-RADS diagnostic categories predicted for breast masses of the *Testing set I* according to histopathology groups.

| Histopathology type | BI-RADS 3 (%) | BI-RADS 4 (%) | BI-RADS 5 (%) |
| --- | --- | --- | --- |
| Fibroadenoma | 2 (1.6) | 19 (15.4) | 0 (0.0) |
| Sclerosing lesions/adenosis | 1 (0.8) | 5 (4.1) | 0 (0.0) |
| Normal breast tissue | 0 (0.0) | 3 (2.4) | 0 (0.0) |
| Inflammatory lesions | 2 (1.6) | 4 (3.3) | 0 (0.0) |
| Papilloma (no atypia) | 2 (1.6) | 4 (3.3) | 0 (0.0) |
| Cysts, ductal ectasia, or seromas | 2 (1.6) | 6 (4.9) | 0 (0.0) |
| Usual ductal hyperplasia | 0 (0.0) | 4 (3.3) | 0 (0.0) |
| Atypical ductal hyperplasia | 3 (2.4) | 5 (4.1) | 0 (0.0) |
| Fibroadenomatoid changes | 0 (0.0) | 1 (0.8) | 0 (0.0) |
| Other benign findings | 1 (0.8) | 8 (6.5) | 0 (0.0) |
| Invasive ductal carcinoma | 1 (0.8) | 40 (32.5) | 0 (0.0) |
| Invasive lobular carcinoma | 0 (0.0) | 3 (2.4) | 0 (0.0) |
| Ductal carcinoma in situ | 0 (0.0) | 2 (1.6) | 0 (0.0) |
| Other malignancies originating from breast tissues | 0 (0.0) | 4 (3.3) | 0 (0.0) |
| Other malignancies (metastases from non breast tissues) | 0 (0.0) | 1 (0.8) | 0 (0.0) |

**Table 7.** Ensemble of support vector machines: BI-RADS diagnostic categories predicted for breast masses of the *Testing set II* according to histopathology groups.

| Histopathology type | BI-RADS 3 (%) | BI-RADS 4 (%) | BI-RADS 5 (%) |
| --- | --- | --- | --- |
| Fibroadenoma | 1 (0.9) | 23 (20.4) | 0 (0.0) |
| Sclerosing lesions/adenosis | 2 (1.8) | 7 (6.2) | 0 (0.0) |
| Normal breast tissue | 0 (0.0) | 2 (1.8) | 0 (0.0) |
| Inflammatory lesions | 1 (0.9) | 1 (0.9) | 0 (0.0) |
| Papilloma (no atypia) | 1 (0.9) | 2 (1.8) | 0 (0.0) |
| Cysts, ductal ectasia, or seromas | 1 (0.9) | 5 (4.4) | 0 (0.0) |
| Usual ductal hyperplasia | 1 (0.9) | 1 (0.9) | 0 (0.0) |
| Atypical ductal hyperplasia | 0 (0.0) | 0 (0.0) | 0 (0.0) |
| Fibroadenomatoid changes | 1 (0.9) | 6 (5.3) | 0 (0.0) |
| Other benign findings | 1 (0.9) | 3 (2.7) | 0 (0.0) |
| Invasive ductal carcinoma | 1 (0.0) | 40 (35.4) | 0 (0.0) |
| Invasive lobular carcinoma | 0 (0.0) | 4 (3.5) | 0 (0.0) |
| Ductal carcinoma in situ | 1 (0.9) | 2 (1.8) | 0 (0.0) |
| Other malignancies originating from breast tissues | 1 (0.9) | 4 (3.5) | 0 (0.0) |
| Other malignancies (metastases from non breast tissues) | 0 (0.0) | 1 (0.9) | 0 (0.0) |

The certified breast radiologist with 34 years of experience in breast imaging accepted the BI-RADS classes of 114 masses (92.7%) and modified the BI-RADS classes of 9 breast masses (7.3%) (Table S5). In 6 of 9 cases the model performed better than the radiologist, since it assigned BI-RADS 3 to masses benign according to histopathology while the radiologist assigned BI-RADS 4. Two breast masses, malignant according to histopathology, were classified by the model as BI-RADS 4 while the radiologist assigned a BI-RADS 5 classification. These masses were invasive ductal carcinomas according to histopathology, thus the radiologist assigned a more appropriate class of malignancy. The last mass was a granular cell tumor at histopathology, usually considered benign, to which mass both the model and the radiologist assigned a wrong malignancy BI-RADS class (BI-RADS 4 and 5, respectively); however we must consider that from a therapeutical point of view this type of tumor (a rare entity derived from Schwann cells) is aggressive and locally recurrent, therefore requiring surgical excision with curative intent [16].

Intra-observer agreement (board-certified breast radiologist with 34 years of experience in breast imaging) in the model classification of BI-RADS was 96% (48/50), with a mean DICE index of 89.7% ± 5.0%. Inter-observer agreement (board-certified breast radiologist with 7 years of experience in breast imaging versus certified breast radiologist with 34 years of experience in breast imaging) in the model classification of BI-RADS was 92% (46/50), with a mean DICE index of 87.0% ± 9.9%.

## 4. Discussion

In this study, we described the development and validation of a radiomic-based machine learning model aimed at predicting the BI-RADS category and reducing the biopsy rate of ultrasound-detected suspicious breast masses, using a series of 821 images of 834 suspicious breast lesions from 819 patients referred to ultrasoundguided core needle biopsy. Of note, the dataset is characterized by a nearly balanced 1.06:1.0 benign-to-malignant ratio according to histopathology, indicating a high level of expertise in lesion selection, already avoiding the biopsy of a large number of benign lesions. The distribution of the histopathology types was expected, considering that lesion selection was based on ultrasound detection, with a very high proportion, among malignancies, of invasive ductal carcinomas (over three quarters), as already reported in similar series [17,18].

Three ensembles of machine learning supervised classifiers were trained using a balanced image set of 299 benign and 299 malignant lesions. The ensemble of support vector machines, based on a qualified majority vote of over 80% for predicting the benign nature of the suspicious masses, showed an over 94% sensitivity (BI-RADS 4–5), allowing to avoid more than 15–18% of biopsies of benign lesions (BI-RADS 3). Interestingly, these performances remained substantially stable when transitioning from internal cross-validation to two external validation sets, with an over 96% sensitivity on images from different ultrasound systems from the same vendor (*Training and internal testing set* and *Testing set I*).

The ability of individual radiomic features to discriminate malignant from benign masses deserve some comments in relation to the classic BI-RADS descriptors [5,19]. This is a crucial point in terms of explainability to radiologists (and patients as well) of the machine learning model output.

The selected radiomic predictors are able to capture shape, margins, and ultrasonographic pattern of suspicious masses consistently with BI-RADS ultrasound descriptors. Morphological predictors such as *compactness* and *acircularity* quantify the deviation of the lesion area from a representative ellipse and circle, respectively, thus being able to distinguish oval and round shape from irregular shapes, the latter more frequent for malignant masses.

The higher values of the c*enter of mass shift* predictor in malignant lesions highlight the more asymmetric spatial distribution of intensities for these lesions. These aspects fit well with findings previously reported by Fleury and Marcomini [20], who noted how lesion shape and margins emerged as the most promising BI-RADS features in distinguishing between benign and malignant lesions.

Several texture predictors showed higher values for malignant than for benign lesions, expressing echo-pattern heterogeneity (captured by different non uniformity features obtained from different texture matrices, i.e., *busyness, zone size non uniformity, grey level non uniformity glszm*, and *dependence count non uniformity*). In addition, the higher values of the texture features *coarseness* and *strength* for benign lesions express the tendency for more homogeneous ultrasonographic textural patterns as indicated by BI-RADS descriptors [5].

Less than 1% of masses were wrongly categorized as BI-RADS 3 in the external *Testing set I*, less than 3% in the external *Testing set II*. Moreover, of the 123 breast lesions of the external *Testing set I*, 114 (92.7%) were categorized in the same class both by the model and the expert radiologist, thus showing the possibility of using the tool as an “expert” second opinion. Of note, considering the nine disagreement cases, the model assigned the correct benign class to six masses, confirming its potential in reducing the biopsy rate of benign masses. The remaining three masses were classified by both the model and the expert radiologist as positive cases, with BI-RADS 4 given by the model, and BI-RADS 5 given by the radiologist, resulting in the same clinical effect, i.e., referral to biopsy. Two were invasive ductal carcinomas, not needing comments. The other was instead a rare entity (a granular cell tumor, usually considered benign but deserving surgery [16]) that can be considered an “expected” false positive case.

It is important to take into consideration the design of this study, that included only ultrasound-detected breast lesions that underwent ultrasound-guided core needle biopsy. In other words, the large number of lesions considered frankly benign at a qualitative observation by the breast radiologists, i.e., those judged to be associated with null likelihood of cancer (BI-RADS 2, mainly being well-circumscribed anechoic cysts or non-modified homogeneously hypoechoic fibroadenomas, both of them with regular margins and frequently also posterior enhancement) did not enter this model training dataset. In addition, this series included both symptomatic and asymptomatic breast masses (as common for consecutive series of ultrasound-detected breast masses in realworld clinical practice), the former having a larger size than the latter. This is mirrored by morphological differences between malignant and benign lesions captured by predictors—such as the *maximum diameter, perimeter* and *area*—found to be larger for the malignant lesions than for benign lesions, reflecting this real-world clinical practice scenario.

In order to validate the clinical utility of our model, its diagnostic performances must be contextualized in the clinical decision making of “to biopsy or not to biopsy” a lesion detected at breast ultrasound. This decision should take into account the high incidence of breast cancer in the female population (in advanced countries, one out of every nine women experiences a breast cancer diagnosis during her lifetime [21–23]) and the increase in cancer probability due to the ultrasound detection of a suspicious lesion, as shown by the experience of targeted ultrasound of mammography-detected [24–26] or MRI-detected lesions [27]. Regarding the inherently high probability of malignancy, we should consider that, in the original consecutive series considered in this work, 451 of 941 lesions (47.9%) were malignant, and that this rate was substantially maintained after technical exclusions due to not sure lesion identification (404 of 834, 48.4%). This context gives a relevant clinical value to the only apparently low specificity (15™18%) provided by our machine learning model, that was still able to maintain an over 94™98% sensitivity. These results measure the potential clinical advantage of the model, meaning the avoidance of about 1 of 6 biopsies of benign lesions even in this selected series (with about 50% of malignancies). Notably, all the machine learning model specificity represents a net gain when compared with the 0% radiologists’ specificity (Table 3), obliged by the study design, including only biopsied lesions.

In a recent work [28], a commercially-available artificial intelligence system based on artificial neural networks was used to evaluate ultrasound-detected breast lesions (classified according BI-RADS categories, from 2 to 5) obtaining a 98% sensitivity, a 97% NPV, and a 64% PPV. Their series was not limited to biopsied lesions only (as was in ours) and the inclusion also of frankly benign lesions (BI-RADS 2) intrinsically increased the specificity of human readers (and of any machine learning model). Indeed, as already observed for diagnostic studies applying breast MRI [29], when considering series solely comprising lesions with histopathology as reference standard the specificity obviously results to be relatively low, because the benign lesions were suspected to be cancer at a degree to deserve biopsy.

This context can also be further understood considering four large-scale series of breast needle biopsies including 3,054 [30], 2,420 [31], 20,001 [32], and 22,297 lesions [33], for a total of 47,772 lesions. The proportions of benign lesions were 54.8%, 44.3%, 51.5%, and 72.6%, respectively, the last series showing that there is no trend in favor of the reduction of the biopsies of benign lesions. Thus, any tool working in this direction is welcome to clinical practice and could be used as a second opinion for clinical decision making in favor of six-month followup (as per the BI-RADS 3 diagnostic category, which was introduced with the aim of avoiding to biopsy too many benign lesions) instead of immediate needle biopsy (as per BI-RADS diagnostic categories 4a or higher [5]). Of course, this possibility, occurring in a real-world clinical scenario, should be sustained by a top-level sensitivity (such as the one achieved by our model) combined with an overall BI-RADS 3 NPV ideally higher than 98%, yielding less than 2% false negatives BI-RADS 3, as recommended by the BI-RADS guidelines [5]. Of note, the NPVs of our model are lower than 98% (78.3%, 92.9%, and 75.0% for the *Training and internal testing set*, the external *Testing set I*, and the external *Testing set II*, respectively) but it regards only on BI-RADS 3 lesions which were all referred to needle biopsy, since our series did not include BI-RADS 3 lesions sent to six-month follow-up. These follow-up cases should have been added to have the overall BI-RADS 3 NPV.

To better clarify the value of our results, we should consider the breast cancer epidemiology at large. According to the International Agency for Research on Cancer [34], in 2020, a total of 2,261,419 new breast cancers were diagnosed worldwide. We can consider that the average rate of benign lesions reported by the four aforementioned large series [30–33] is 29,235 of 47,772 (61.1%), rounded to 60% (meaning a 40% malignancy rate), and that the majority of breast needle biopsy are performed under ultrasound guidance (with at least a 70% estimate [2,17,35,36]). Even applying a tool providing only a 15% additional specificity we could already save about 356,000 biopsies, i.e., 15% of the 2,375,000 needle biopsies of benign lesions performed worldwide under ultrasound guidance every year.

The value of our tool could be much greater when used in conjunction with the physician’s evaluation. There are indeed already some studies that demonstrate an increase in physician performance when the decision whether to perform a biopsy or refer to follow-up is made with the support of decision systems based on AI models predicting the risk of malignancy of a lesion. For example, in the experience reported by Zhao et al. [37], the feasibility of a deep learning-based computer-aided diagnosis (CAD) system was explored in order to improve the diagnostic accuracy of residents in detecting BI-RADS 4a lesions. The authors evaluated the improvement obtained by downgrading BI-RADS 4a lesions identified by radiologists to possibly benign lesions as per CAD prediction. The sensitivity of the integrated results remained at a relatively high level (> 92.7%), while the specificities of all residents significantly improved after using the results of CAD, rising from 19.5–48.7% to 46.0–76.1%. Similarly, Barinov et al. [38] showed that through simple fusion schemes they could increase performance beyond that of either their CAD system or the radiologist alone, obtaining an absolute average PPV increase of ∼15% while keeping original radiologists’ sensitivity. Especially less-experienced radiologists could benefit from a CAD system for the diagnosis of ultrasound-detected breast masses, as shown by Lee et al. [39] who compared the evaluation of 500 lesions performed by five experienced and five inexperienced radiologists, with and without CAD; the diagnostic performance of the inexperienced group after combination with CAD result was significantly improved (ROC-AUC 0.71, 95% CI 0.65–0.77) compared with the diagnostic performance without CAD (ROC-AUC 0.65, 95% CI 0.58–0.71).

However, we should also consider that the final decision to biopsy or to follow-up an ultrasound-detect breast lesion also depends on factors other than ultrasound image characteristics, i.e. on family and personal history of the patient (including the absence of presence of symptoms), and the possible preceding lesion detection on other imaging techniques such as mammography/tomosynthesis or MRI. In this study we did not take into consideration these different indications to breast ultrasound. In addition, also the patient’s psychological condition plays a relevant role in the final decision making. From this viewpoint, the improvement of clinical decision making that can be obtained using our model could be estimated in a prospective clinical study and/or in a retrospective reader study, where BI-RADS classes are assigned by our model (based on the consensus of votes expressed by the support vector classifiers of the best ensemble) and then proposed to physicians (e.g. the highest consensus for malignancy leads to the highest likelihood of cancer, i.e., BI-RADS 5). Regarding the role of the BI-RADS 3 category in this study, we highlight that here we considered only lesions that undergo needle biopsy, not those that were sent to follow-up and finally downgraded to BI-RADS 2 (for example for reduction in size), with no possibility to obtain histopathology reference standard. Hence, the potential benefit of the AI tool system could be explored in followed-up lesions with final benign outcome, to assess the role of the model in this specific setting.

A limitation of this study is related to the origin of its patient cohort (a University Hospital located in Northern Italy), which is therefore composed of lesions observed in European Caucasian subjects. While the ultrasound appearance of benign and malignant lesions should not be different in other ethnicities, the different structure of the breast (e.g., Asian women have breasts denser than Caucasian women [40–42]) could influence the relation between the lesion and the surrounding tissue: an isoechoic lesion surrounded by fat may be hypoechoic is surrounded by gland parenchyma. However, considering that our model takes into consideration absolute and not relative signal intensities, we do not expect different performances.

In conclusion, in this study, a specifically-developed machine learning model based on radiomics to predict the malignant or benign nature of ultrasound-detected suspicious breast lesions was first trained and cross-validated on 598 images of pathology-proven benign or malignant lesions, then underwent independent external validation on 236 other images. Such a model was proven to be effective in predicting BI-RADS 3, 4, and 5 classes and potentially clinically useful in providing an over 15% reduction of the biopsy rate of lesions finally revealed as benign, while still warranting very high sensitivity. This system can be used in a clinical context as a decision support system to support radiologists in the assignment of BI-RADS classes and toward decision of short-interval follow-up versus tissue sampling for suspicious breast lesions.

## Data Availability

All data analysed for this study are presented in the manuscript or in the supplementary material.

## Supplementary Materials

**Table S1**: Ensemble of random forest classifiers. Classification performance and statistical significance with respect to chance/random classification (*p* value). Performances are reported for a majority vote of 50% and for the internal testing, **Table S2**: Ensembles of support vector machine classifiers. Classification performances and statistical significance with respect to chance/random classification (*p* value). Performances are reported for a majority vote of 50% and for the internal testing, **Table S3**: Ensembles of k nearest neighbors classifiers. Classification performances and statistical significance with respect to chance/random classification (*p* value). Performances are reported for a majority vote of 50% and for the internal testing, **Table S4**: Complete list of 107 radiomic features with the values of the four representative lesions (two benign and two malignant) shown in Figures 3 and 4, **Table S5**: BI-RADS classes assigned by the ensemble of support vector machines (AI model) and the certified breast radiologist, **Figure S1**: Violin and box plots of the most relevant features ranked from 16 to 25.

## Author Contributions

Conceptualization, C.S., I.C. and F.S.; Data curation, M.I., C.S., V.M., G.C., E.S., A.C., S.S. and L.A.C.; Formal analysis, M.I., C.S., E.S. and I.C.; Funding acquisition, C.S., I.C. and F.S.; Investigation, M.I., C.S., V.M., G.C., E.S., A.C., S.S. and L.A.C.; Methodology, M.I., C.S., A.C., I.C. and F.S.; Project administration, C.S. and I.C.; Resources, C.S., V.M., E.S., S.S., L.A.C., I.C. and F.S.; Software, M.I., C.S., G.C., E.S. and I.C.; Supervision, S.S., L.A.C., I.C. and F.S.; Validation, M.I., C.S., V.M., A.C., I.C. and F.S.; Visualization, M.I., G.C., E.S. and A.C.; Writing – original draft, M.I., G.C., I.C. and F.S.; Writing – review & editing, M.I., C.S., V.M., G.C., E.S., A.C., S.S., L.A.C., I.C. and F.S.

## Institutional Review Board Statement

The study was conducted according to the guidelines of the Declaration of Helsinki, and approved by the Ethics Committee of IRCCS Ospedale San Raffaele (protocol code “SenoRetro”, first approved on November 9th, 2017, then amended on July 18th, 2019, and on May 12th, 2021).

## Informed Consent Statement

Patient consent was waived due to the retrospective nature of the study.

## Data Availability Statement

All data analysed for this study are presented in the manuscript or in the supplementary material.

