## Supplementary material for "A Machine Learning Ensemble Based on Radiomics to Predict BI-RADS Category and Reduce the Biopsy Rate of Ultrasound-Detected Suspicious Breast Masses"

**Table S1.** Ensemble of random forest classifiers. Classification performance and statistical significance with respect to chance/random classification (*p* value). Performances are reported for a majority vote of 50% and for the internal testing.

| Metric | Internal testing |
|---|---|
| ROC-AUC (%) (95% confidence interval) | 68** (67–69) |
| Sensitivity (%) (95% confidence interval) | 64** (60–68) |
| Specificity (%) (95% confidence interval) | 63** (62–64) |
| Positive predictive value (%) (95% confidence interval) | 63** (62–64) |
| Negative predictive value (%) (95% confidence interval) | 63** (62–65) |

** denotes a statistical significance at 0.005 (adjusted with Bonferroni-Holm correction).

**Table S2.** Ensembles of support vector machine classifiers. Classification performances and statistical significance with respect to chance/random classification (*p* value). Performances are reported for a majority vote of 50% and for the internal testing.

| Metric | Internal testing |
|---|---|
| ROC-AUC (%) (95% confidence interval) | 75** (74–75) |
| Sensitivity (%) (95% confidence interval) | 72** (71–73) |
| Specificity (%) (95% confidence interval) | 70** (66–74) |
| Positive predictive value (%) (95% confidence interval) | 71** (67–74) |
| Negative predictive value (%) (95% confidence interval) | 71** (70–73) |

** denotes a statistical significance at 0.005 (adjusted with Bonferroni-Holm correction).

**Table S3.** Ensembles of k nearest neighbors classifiers. Classification performances and statistical significance with respect to chance/random classification (*p* value). Performances are reported for a majority vote of 50% and for the internal testing.

| Metric | Internal testing |
|---|---|
| ROC-AUC (%) (95% confidence interval) | 74** (73–75) |
| Sensitivity (%) (95% confidence interval) | 70** (67–72) |
| Specificity (%) (95% confidence interval) | 67** (61–73) |
| Positive predictive value (%) (95% confidence interval) | 68** (64–72) |
| Negative predictive value (%) (95% confidence interval) | 69** (67–71) |

** denotes a statistical significance at 0.005 (adjusted with Bonferroni-Holm correction).

**Table S4.** Complete list of 107 radiomic features with the values of the four representative lesions (two benign and two malignant) shown in Figures 3 and 4.

| Feature family | Feature name | Unit | Figure 3a (benign) | Figure 3b (benign) | Figure 4a (malignant) | Figure 4b (malignant) |
|---|---|---|---|---|---|---|
| Morphology | Area | $mm^2$ | 37.489 | 143.634 | 214.474 | 60.387 |
| Morphology | Perimeter | mm | 24.165 | 50.155 | 68.826 | 35.145 |
| Morphology | Perimeter to area ratio | mm | 0.645 | 0.349 | 0.321 | 0.582 |
| Morphology | Compactness | – | 0.807 | 0.718 | 0.569 | 0.614 |
| Morphology | Circularity | – | 0.898 | 0.847 | 0.754 | 0.784 |
| Morphology | Acircularity | – | 0.113 | 0.181 | 0.326 | 0.276 |
| Morphology | Center of mass shift | mm | 3.495 | 4.458 | 3.997 | 5.861 |
| Morphology | Maximum diameter | mm | 8.934 | 16.264 | 21.153 | 13.951 |
| Intensity-based statistics | Mean | – | 11.369 | 62.602 | 29.547 | 67.277 |
| Intensity-based statistics | Variance | – | 140.133 | 426.847 | 500.680 | 364.277 |
| Intensity-based statistics | Median | – | 7 | 60 | 25 | 66 |
| Intensity-based statistics | Minimum | – | 0 | 10 | 0 | 15 |
| Intensity-based statistics | $10^{th}$ percentile | – | 2 | 38 | 5 | 44 |
| Intensity-based statistics | $90^{th}$ percentile | – | 27 | 89 | 60 | 90 |
| Intensity-based statistics | Interquartile range | – | 10 | 26 | 29 | 23 |
| Intensity-based statistics | Mean absolute deviation | – | 8.2323 | 15.9370 | 17.5030 | 14.4264 |
| Intensity-based statistics | Robust mean absolute deviation | – | 4.7193 | 10.9774 | 12.5114 | 9.5833 |

| | | | | | | |
|---|---|---|---|---|---|---|
| Intensity-based statistics | Median absolute deviation | – | 7.4631 | 15.8155 | 17.1981 | 14.4122 |
| Intensity-based statistics | Coefficient of variation | – | 1.0412 | 0.3300 | 0.7573 | 0.2837 |
| Intensity-based statistics | Quartile coefficient | – | 0.5556 | 0.2131 | 0.5472 | 0.1729 |
| Intensity-based statistics | Energy | – | 869080 | 53714028 | 25352896 | 49809980 |
| Intensity-based statistics | Root mean | – | 16.413 | 65.923 | 37.063 | 69.932 |
| Intensity histogram | Mean | – | 9.675 | 17.789 | 11.821 | 23.116 |
| Intensity histogram | Variance | – | 89.795 | 45.926 | 73.689 | 68.073 |
| Intensity histogram | Median | – | 6 | 17 | 10 | 23 |
| Intensity histogram | 10th percentile | – | 2 | 10 | 2 | 13 |
| Intensity histogram | 90th percentile | – | 22 | 26 | 23 | 33 |
| Intensity histogram | Maximum | – | 64 | 64 | 64 | 64 |
| Intensity histogram | Range | – | 63 | 63 | 63 | 63 |
| Intensity histogram | Mean absolute deviation | – | 6.576 | 5.226 | 6.713 | 6.214 |
| Intensity histogram | Robust mean absolute deviation | – | 3.773 | 3.600 | 4.980 | 4.241 |
| Intensity histogram | Median absolute deviation | – | 5.983 | 5.183 | 6.583 | 6.201 |
| Intensity histogram | Coefficient of variation | – | 0.979 | 0.381 | 0.726 | 0.357 |
| Intensity histogram | Entropy | – | 4.494 | 4.738 | 4.806 | 5.004 |
| Intensity histogram | Uniformity | – | 0.065 | 0.045 | 0.044 | 0.039 |
| Intensity histogram | Minimum histogram gradient | – | -157.5 | -134.5 | -332.0 | -175.5 |

| | | | | | | |
|---|---|---|---|---|---|---|
| Grey-level co-occurrence matrix | Joint maximum | – | 0.040 | 0.013 | 0.047 | 0.015 |
| Grey-level co-occurrence matrix | Joint average | – | 9.121 | 17.690 | 11.684 | 22.991 |
| Grey-level co-occurrence matrix | Joint variance | – | 75.650 | 44.367 | 71.977 | 66.033 |
| Grey-level co-occurrence matrix | Joint entropy | – | 7.830 | 8.218 | 7.879 | 8.619 |
| Grey-level co-occurrence matrix | Difference average | – | 2.843 | 2.217 | 1.858 | 2.473 |
| Grey-level co-occurrence matrix | Difference variance | – | 11.931 | 4.380 | 3.717 | 5.181 |
| Grey-level co-occurrence matrix | Difference entropy | – | 3.138 | 2.765 | 2.606 | 2.906 |
| Grey-level co-occurrence matrix | Sum average | – | 18.242 | 35.381 | 23.369 | 45.982 |
| Grey-level co-occurrence matrix | Sum variance | – | 282.587 | 168.172 | 280.738 | 252.834 |
| Grey-level co-occurrence matrix | Sum entropy | – | 5.420 | 5.686 | 5.807 | 5.978 |
| Grey-level co-occurrence matrix | Angular second moment | – | 0.0106 | 0.0051 | 0.0086 | 0.0041 |
| Grey-level co-occurrence matrix | Contrast | – | 20.012 | 9.294 | 7.169 | 11.297 |
| Grey-level co-occurrence matrix | Dissimilarity | – | 2.843 | 2.217 | 1.858 | 2.473 |
| Grey-level co-occurrence matrix | Inverse difference | – | 0.444 | 0.449 | 0.508 | 0.426 |
| Grey-level co-occurrence matrix | Inverse difference normalised | – | 0.960 | 0.967 | 0.973 | 0.964 |
| Grey-level co-occurrence matrix | Inverse difference moment | – | 0.373 | 0.377 | 0.446 | 0.349 |
| Grey-level co-occurrence matrix | Inverse difference moment normalised | – | 0.995 | 0.998 | 0.998 | 0.997 |
| Grey-level co-occurrence matrix | Inverse variance | – | 0.312 | 0.364 | 0.370 | 0.340 |
| Grey-level co-occurrence matrix | Autocorrelation | – | 148.839 | 352.667 | 204.915 | 588.976 |

| | | | | | | |
|---|---|---|---|---|---|---|
| Grey-level co-occurrence matrix | Cluster tendency | – | 282.587 | 168.172 | 280.738 | 252.834 |
| Grey-level co-occurrence matrix | First measure of information correlation | – | -0.223 | -0.259 | -0.354 | -0.271 |
| Grey-level run length matrix | Short run emphasis | – | 0.874 | 0.879 | 0.841 | 0.894 |
| Grey-level run length matrix | Long runs emphasis | – | 1.966 | 1.735 | 2.379 | 1.668 |
| Grey-level run length matrix | Low grey level run emphasis | – | 0.0745 | 0.0060 | 0.0465 | 0.0038 |
| Grey-level run length matrix | High grey level run emphasis | – | 212.347 | 371.377 | 242.140 | 615.364 |
| Grey-level run length matrix | Short run low grey level emphasis | – | 0.0525 | 0.0052 | 0.0295 | 0.0033 |
| Grey-level run length matrix | Short run high grey level emphasis | – | 203.341 | 332.728 | 219.252 | 558.174 |
| Grey-level run length matrix | Long run low grey level emphasis | – | 0.312 | 0.011 | 0.307 | 0.007 |
| Grey-level run length matrix | Long run high grey level emphasis | – | 260.514 | 603.440 | 375.931 | 961.905 |
| Grey-level run length matrix | Grey level non uniformity | – | 608.175 | 1824.154 | 2270.620 | 1327.523 |
| Grey-level run length matrix | Grey level non uniformity normalized | – | 0.0581 | 0.0443 | 0.0402 | 0.0384 |
| Grey-level run length matrix | Run length non uniformity | – | 7548.2 | 30082.6 | 37346.9 | 26255.1 |
| Grey-level run length matrix | Run length non uniformity normalized | – | 0.722 | 0.731 | 0.662 | 0.759 |
| Grey-level run length matrix | Run percentage | – | 0.811 | 0.833 | 0.764 | 0.849 |
| Grey-level run length matrix | Grey level variance | – | 99.103 | 47.546 | 75.238 | 69.208 |
| Grey-level run length matrix | Run length variance | – | 0.445 | 0.294 | 0.668 | 0.280 |
| Grey-level run length matrix | Run entropy | – | 5.395 | 5.528 | 5.829 | 5.720 |
| Grey-level size zone matrix | Small zone emphasis | – | 0.718 | 0.636 | 0.625 | 0.672 |

| | | | | | | |
|---|---|---|---|---|---|---|
| Grey-level size zone matrix | Large zone emphasis | – | 21.219 | 8.482 | 269.335 | 7.599 |
| Grey-level size zone matrix | Low grey level zone emphasis | – | 0.0381 | 0.0056 | 0.0196 | 0.0035 |
| Grey-level size zone matrix | High grey level zone emphasis | – | 299.7 | 401.7 | 313.5 | 657.7 |
| Grey-level size zone matrix | Small zone low grey level emphasis | – | 0.0192 | 0.0032 | 0.0086 | 0.0021 |
| Grey-level size zone matrix | Small zone high grey level emphasis | – | 260.663 | 273.737 | 223.044 | 465.417 |
| Grey-level size zone matrix | Large zone high grey level emphasis | – | 7.183 | 0.056 | 67.193 | 0.035 |
| Grey-level size zone matrix | Grey level non uniformity glszm | – | 69.409 | 252.892 | 259.508 | 194.439 |
| Grey-level size zone matrix | Grey level non uniformity normalized glszm | – | 0.0445 | 0.0425 | 0.0364 | 0.0369 |
| Grey-level size zone matrix | Zone size non uniformity | – | 743.6 | 2255.5 | 2600.2 | 2209.0 |
| Grey-level size zone matrix | Zone size non uniformity normalized | – | 0.477 | 0.379 | 0.365 | 0.420 |
| Grey-level size zone matrix | Zone percentage glszm | – | 0.484 | 0.482 | 0.386 | 0.517 |
| Grey-level size zone matrix | Grey level variance glszm | – | 122.985 | 52.887 | 79.570 | 73.058 |
| Grey-level size zone matrix | Zone size variance | – | 16.943 | 4.170 | 262.637 | 3.854 |
| Grey-level size zone matrix | Zone size entropy | – | 6.407 | 6.750 | 7.036 | 6.820 |
| Neighbourhood grey tone difference matrix | Coarseness | – | 0.0037 | 0.0015 | 0.0013 | 0.0022 |
| Neighbourhood grey tone difference matrix | Contrast | – | 0.0923 | 0.0390 | 0.0475 | 0.0410 |
| Neighbourhood grey tone difference matrix | Busyness | – | 0.521 | 0.545 | 1.052 | 0.243 |
| Neighbourhood grey tone difference matrix | Strength | – | 17.164 | 3.142 | 4.050 | 5.143 |
| Neighbouring grey level dependence matrix | Low dependence emphasis | – | 0.421 | 0.394 | 0.324 | 0.432 |

| | | | | | | |
|---|---|---|---|---|---|---|
| Neighbouring grey level dependence matrix | High dependence emphasis | – | 8.809 | 6.908 | 11.245 | 6.241 |
| Neighbouring grey level dependence matrix | Low grey level count emphasis | – | 0.100 | 0.006 | 0.070 | 0.004 |
| Neighbouring grey level dependence matrix | High grey level count emphasis | – | 183.404 | 362.360 | 213.429 | 602.416 |
| Neighbouring grey level dependence matrix | Low dependence low grey level emphasis | – | 0.0163 | 0.0021 | 0.0075 | 0.0014 |
| Neighbouring grey level dependence matrix | Low dependence high grey level emphasis | – | 133.879 | 161.772 | 103.073 | 288.651 |
| Neighbouring grey level dependence matrix | High dependence low grey level emphasis | – | 2.165 | 0.044 | 2.032 | 0.026 |
| Neighbouring grey level dependence matrix | Grey level non uniformity | – | 211.159 | 555.511 | 812.533 | 397.161 |
| Neighbouring grey level dependence matrix | Grey level non uniformity normalized | – | 0.0655 | 0.0449 | 0.0440 | 0.0390 |
| Neighbouring grey level dependence matrix | Dependence count non uniformity | – | 737.2 | 3177.8 | 3651.0 | 2777.8 |
| Neighbouring grey level dependence matrix | Dependence count non uniformity normalized | – | 0.229 | 0.257 | 0.198 | 0.273 |
| Neighbouring grey level dependence matrix | Grey level variance | – | 89.795 | 45.926 | 73.689 | 68.073 |
| Neighbouring grey level dependence matrix | Dependence count variance | – | 2.486 | 1.446 | 2.925 | 1.354 |
| Neighbouring grey level dependence matrix | Dependence count entropy | – | 6.473 | 6.831 | 7.030 | 6.990 |
| Neighbouring grey level dependence matrix | Dependence count energy | – | 0.0158 | 0.0115 | 0.0098 | 0.0109 |

**Table S5**. BI-RADS classes assigned by the ensemble of support vector machines (AI model) and the certified breast radiologist

| AI model | Certified breast radiologist |
|---|---|
| BI-RADS 3 | BI-RADS 4 |
| BI-RADS 4 | BI-RADS 5 |
| BI-RADS 4 | BI-RADS 4 |
| BI-RADS 4 | BI-RADS 4 |
| BI-RADS 4 | BI-RADS 4 |
| BI-RADS 4 | BI-RADS 4 |
| BI-RADS 4 | BI-RADS 4 |
| BI-RADS 4 | BI-RADS 4 |
| BI-RADS 4 | BI-RADS 4 |
| BI-RADS 3 | BI-RADS 3 |
| BI-RADS 3 | BI-RADS 4 |
| BI-RADS 4 | BI-RADS 4 |
| BI-RADS 4 | BI-RADS 4 |
| BI-RADS 3 | BI-RADS 4 |
| BI-RADS 3 | BI-RADS 3 |
| BI-RADS 3 | BI-RADS 4 |
| BI-RADS 4 | BI-RADS 5 |
| BI-RADS 4 | BI-RADS 4 |
| BI-RADS 3 | BI-RADS 3 |
| BI-RADS 4 | BI-RADS 4 |
| BI-RADS 4 | BI-RADS 4 |
| BI-RADS 4 | BI-RADS 4 |
| BI-RADS 4 | BI-RADS 4 |
| BI-RADS 4 | BI-RADS 4 |
| BI-RADS 4 | BI-RADS 4 |
| BI-RADS 4 | BI-RADS 4 |
| BI-RADS 4 | BI-RADS 4 |
| BI-RADS 4 | BI-RADS 4 |
| BI-RADS 4 | BI-RADS 4 |
| BI-RADS 4 | BI-RADS 4 |

| | |
|---|---|
| BI-RADS 4 | BI-RADS 4 |
| BI-RADS 4 | BI-RADS 4 |
| BI-RADS 4 | BI-RADS 4 |
| BI-RADS 4 | BI-RADS 4 |
| BI-RADS 4 | BI-RADS 4 |
| BI-RADS 3 | BI-RADS 3 |
| BI-RADS 4 | BI-RADS 4 |
| BI-RADS 4 | BI-RADS 4 |
| BI-RADS 4 | BI-RADS 4 |
| BI-RADS 4 | BI-RADS 4 |
| BI-RADS 4 | BI-RADS 4 |
| BI-RADS 4 | BI-RADS 4 |
| BI-RADS 3 | BI-RADS 3 |
| BI-RADS 4 | BI-RADS 4 |
| BI-RADS 4 | BI-RADS 4 |
| BI-RADS 4 | BI-RADS 4 |
| BI-RADS 4 | BI-RADS 4 |
| BI-RADS 4 | BI-RADS 4 |
| BI-RADS 4 | BI-RADS 4 |
| BI-RADS 4 | BI-RADS 4 |
| BI-RADS 4 | BI-RADS 4 |
| BI-RADS 4 | BI-RADS 4 |
| BI-RADS 4 | BI-RADS 4 |
| BI-RADS 4 | BI-RADS 4 |
| BI-RADS 4 | BI-RADS 4 |
| BI-RADS 4 | BI-RADS 4 |
| BI-RADS 4 | BI-RADS 4 |
| BI-RADS 4 | BI-RADS 4 |
| BI-RADS 4 | BI-RADS 4 |
| BI-RADS 4 | BI-RADS 4 |
| BI-RADS 4 | BI-RADS 5 |
| BI-RADS 4 | BI-RADS 4 |
| BI-RADS 4 | BI-RADS 4 |
| BI-RADS 4 | BI-RADS 4 |

| | |
|---|---|
| BI-RADS 4 | BI-RADS 4 |
| BI-RADS 4 | BI-RADS 4 |
| BI-RADS 4 | BI-RADS 4 |
| BI-RADS 4 | BI-RADS 4 |
| BI-RADS 3 | BI-RADS 3 |
| BI-RADS 4 | BI-RADS 4 |
| BI-RADS 4 | BI-RADS 4 |
| BI-RADS 4 | BI-RADS 4 |
| BI-RADS 4 | BI-RADS 4 |
| BI-RADS 4 | BI-RADS 4 |
| BI-RADS 4 | BI-RADS 4 |
| BI-RADS 4 | BI-RADS 4 |
| BI-RADS 4 | BI-RADS 4 |
| BI-RADS 4 | BI-RADS 4 |
| BI-RADS 4 | BI-RADS 4 |
| BI-RADS 4 | BI-RADS 4 |
| BI-RADS 4 | BI-RADS 4 |
| BI-RADS 4 | BI-RADS 4 |
| BI-RADS 4 | BI-RADS 4 |
| BI-RADS 4 | BI-RADS 4 |
| BI-RADS 4 | BI-RADS 4 |
| BI-RADS 4 | BI-RADS 4 |
| BI-RADS 4 | BI-RADS 4 |
| BI-RADS 4 | BI-RADS 4 |
| BI-RADS 4 | BI-RADS 4 |
| BI-RADS 4 | BI-RADS 4 |
| BI-RADS 4 | BI-RADS 4 |
| BI-RADS 4 | BI-RADS 4 |
| BI-RADS 4 | BI-RADS 4 |
| BI-RADS 4 | BI-RADS 4 |
| BI-RADS 4 | BI-RADS 4 |
| BI-RADS 4 | BI-RADS 4 |
| BI-RADS 4 | BI-RADS 4 |
| BI-RADS 4 | BI-RADS 4 |

| | |
|---|---|
| BI-RADS 4 | BI-RADS 4 |
| BI-RADS 3 | BI-RADS 4 |
| BI-RADS 3 | BI-RADS 4 |
| BI-RADS 4 | BI-RADS 4 |
| BI-RADS 4 | BI-RADS 4 |
| BI-RADS 4 | BI-RADS 4 |
| BI-RADS 4 | BI-RADS 4 |
| BI-RADS 4 | BI-RADS 4 |
| BI-RADS 4 | BI-RADS 4 |
| BI-RADS 3 | BI-RADS 3 |
| BI-RADS 3 | BI-RADS 3 |
| BI-RADS 4 | BI-RADS 4 |
| BI-RADS 4 | BI-RADS 4 |
| BI-RADS 4 | BI-RADS 4 |
| BI-RADS 4 | BI-RADS 4 |
| BI-RADS 4 | BI-RADS 4 |
| BI-RADS 4 | BI-RADS 4 |
| BI-RADS 4 | BI-RADS 4 |
| BI-RADS 4 | BI-RADS 4 |
| BI-RADS 4 | BI-RADS 4 |
| BI-RADS 4 | BI-RADS 4 |
| BI-RADS 4 | BI-RADS 4 |
| BI-RADS 4 | BI-RADS 4 |
| BI-RADS 4 | BI-RADS 4 |
| BI-RADS 4 | BI-RADS 4 |

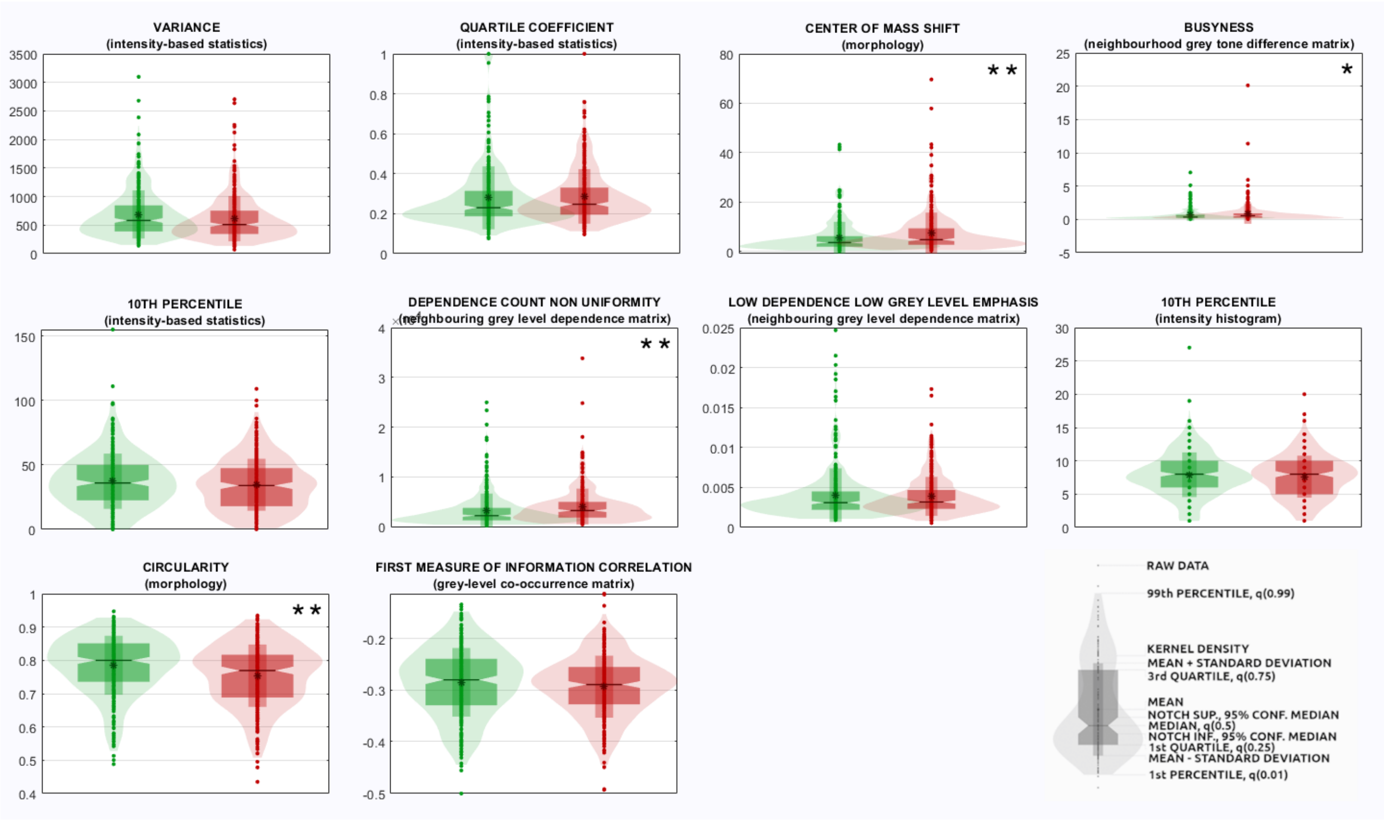


**Figure S1.** Violin and box plots of the most relevant features ranked from 16 to 25

Green: benign class. Red: malignant class.

* denotes statistical significance at 0.05 (adjusted with Bonferroni-Holm correction). ** denotes a statistical significance at 0.005 (adjusted with Bonferroni-Holm correction).
